# Pre-Post Analysis of the Impact of British Columbia Nurse Practitioner Primary Care Clinics on Patient Health and Care Experience

**DOI:** 10.1101/2023.02.13.23285874

**Authors:** Damien Contandriopoulos, Katherine Bertoni, Arnaud Duhoux, Gurprit K. Randhawa

## Abstract

**Objective:** This study aims to evaluate the impact of a primary care nurse-practitioner-led clinic model piloted in British Columbia (Canada) on patients’ health and care experience.

**Design:** The study relies on a quasi-experimental longitudinal design based on a pre-and-post survey of patients receiving care in NP-Led clinics. The pre-rostering survey (T0) was focused on patients’ health status and care experiences preceding being rostered to the NP clinic. One year later, patients were asked to complete a similar survey (T1) focused on the care experiences with the NP clinic.

**Setting:** To solve recurring problems related to poor primary care accessibility, British Columbia opened four pilot NP-led clinics in 2020. Each clinic has the equivalent of approximately six full-time NPs, four other clinicians plus support staff. Clinics are located in four cities ranging from core urban to peri rural.

**Participants:** Recruitment was conducted by the clinic’s clerical staff or by their care provider. A total of 437 usable T0 surveys and 254 matched and usable T1 surveys were collected.

**Primary outcome measures:** The survey instrument was focused on five core dimensions of patients’ primary care experience (accessibility, continuity, comprehensiveness, responsiveness, and outcomes of care) as well as on the SF-12 Short-form Health Survey.

**Results:** Scores for all dimensions of patients’ primary care experience increased significantly: Accessibility (T0=5.9, T1=7.9, p<0.000), Continuity (T0=5.5, T1=8.8, p<0.000), Comprehensiveness (T0=5.6, T1=8.4, p<0.000), Responsiveness (T0=7.2, T1=9.5, p<0.000), Outcomes of care (T0=5.0, T1=8.3, p<0.000). SF-12 Physical health T-scores also rose significantly (T0=44.8, T1=47.6, p<0.000) but no changes we found in the mental health T scores (T0=45.8, T1=46.3 p=0.709).

**Conclusions:** Our results suggest that the NP-Led primary care model studied here likely constitutes an effective approach to improve primary care accessibility and quality.

**Strengths and limitations:**

- This study evaluates the impact of a primary care nurse-practitioner-led clinic model piloted in British Columbia (Canada) on patients’ health and care experience
- The study rests on a pre-post survey without a control group therefore the differences observed could be caused by external factors
- Data collection took place between 2020 and 2022, during the Covid-19 pandemic.
- Only four NP-PCC clinics exist and participation in the survey was voluntary and uncompensated limiting the number of respondents

## INTRODUCTION

The lack of timely access to primary care services has been a significant problem across Canada for decades. Nearly one in seven Canadians had reported not having a regular healthcare provider in 2019 (1) and, in 2020, 40% reported having an avoidable emergency department visit because of the lack of primary care options (2). Available indicators also suggest the situation has significantly worsened since the Covid-19 pandemic.

Increased reliance on non-physicians such as registered nurses and Nurse Practitioners (NPs) has long been documented as a way to improve primary care access (3, 4, 5, 6, 7, 8). In Canada, NPs are registered nurses with strong practice experience as well as a specialized master or doctorate level training. They have a broader scope of practice including the ability to autonomously diagnose, prescribe treatments and refer patients to test or specialist physicians.

Several models have been used to integrate NPs into primary care delivery. The model with the highest disruption potential is called the NP-Led Clinic (9). Those clinics have NPs as the pivot professional of a team providing the full scope of primary care services to a panel of patients (3, 10). The NP-led clinic model has consistently been found to be effective and efficient in meeting the primary care needs of a general population (6, 8, 11, 12, 13).

Despite the strength and abundance of evidence supporting the NP-Led model, its implementation in Canada has been limited and patchy. In 2020, in response to the widespread need for primary care and the successful advocacy from its NP association (14), British Columbia (BC), one of Canada’s 10 provinces, opened four pilot NP-led clinics. The BC model was named Nurse-Practitioner Primary Care Clinic (NP-PCC).

The aim of this study is to evaluate the impact of NP-PCCs on the health and patient reported experience of the first cohorts of rostered patients.

## BACKGROUND

The pilot NP-PCCs are located in four cities ranging from core urban to peri-rural. The Axis clinic is in Surrey, Nexus is in Nanaimo, Health Care on Yates (HCOY) is in Victoria and Flowerstone is in Qualicum Beach. From the onset, the clinics were tasked with providing comprehensive primary care services to people from formally defined catchment areas who were without a primary care provider. The rostering goal for all clinics was set at 6800 patients after 3 years of operation by the Ministry of Health. Patients are formally rostered to the clinic as well as informally attached to a specific NP within the clinic. Due to very high demand, all four clinics used patient waitlists or random draws as a part of their attachment processes. On average, each clinic has the equivalent of six full-time NPs, one social worker, one mental health counsellor or clinician, two RNs, one clinic manager, and four Medical Office Assistants. Clinics are funded by the BC Ministry of Health and have a self-appointed board of directors and managerial autonomy (15).

## DATA AND METHODS

### Study design

The study relies on a quasi-experimental, longitudinal design based on a pre-and-post survey of patients receiving care in a NP-PCC. The pre-rostering survey (Time zero or T0) asked respondents to describe their care experience during the two years preceding being rostered to a NP-PCC as well as their current health. Approximately one year later, patients were asked to complete a second (Time one or T1) similar survey, this time focused on their care experience as an NP-PCC patient. The design was adapted from previous work done by the same team (10, 16, 17).

### Participant Recruitment

For the T0 recruitment, patients newly rostered to one of the clinics were given an information sheet about the project by the clinic’s clerical staff or by their care provider. The information sheet included a link to a consent form and online survey instrument hosted on the University of Victoria Survey Monkey platform. The survey asked patients to provide their government-issued personal health number (PHN). The PHNs of patients who completed the T0 survey were shared with the clinics’ clerical staff who made a note in the clinics’ Electronic Health Record system. About a year later during a subsequent visits to the clinic, those patients were provided with an invitation to complete the T1 survey. Email invitations were also sent by the clinics to patients identified as having completed the T0 survey. Data collection periods varied slightly from site to site (see Figure 5 in Appendix) but, overall, the T0 period ran from November 2020 to January 2022 and the T1 period ran from December 2021 to December 2022. The study was approved by the University of Victoria Human Research Ethics Board (#20-0324)

### Instruments

The patient reported experience from being rostered in a NP-PCC were measured according to five core dimensions of primary care experience: accessibility, continuity, comprehensiveness, responsiveness, and outcomes of care. Those dimensions have been extensively discussed and conceptualized in the literature (18, 19, 20). The definition of accessibility rests on Donabedian’s seminal work (21), as the fit between the structures of production, on one hand, and society’s needs and their geographic distribution, on the other. It is operationalized here as the patient’s perception that they can access healthcare services without undue constraints. Continuity is defined, based on the work of Haggerty et al. (22), as the fact that a patient is treated by the same professional or the same team over time (relational continuity) and that different services are harmoniously integrated with each other (management continuity). Comprehensiveness encompasses two dimensions that make up the scope of patient management: considering all of a patient’s needs and providing a complete basket of services (10, 23). Responsiveness is defined here as the convergence between the patients’ expectations regarding non-technical elements of the care and what the clinic offers in practice (20). Finally, what is described as outcomes of care relates to the patients’ perception that the care received had a positive impact on their health (24).

The five dimensions described above were measured using a survey instrument adapted from the one developed by Pineault et al. (24, 25) which was in turn an expanded version of two well-validated pre-existing instruments: the Primary Care Assessment Survey (26) and the Primary Care Assessment Tool (27). Details on the phrasing of the survey questions and computation of scores can be found elsewhere (17, 25). Each score is measured on a scale ranging from 0 to 10.

In addition to the patient-focused indicators described above, we also measured respondents’ general health status using the SF-12 Short-form Health Survey (Ware et al., 1996). The SF-12 instrument provides a physical health score and a mental health score, both measured on a scale ranging from 0 to 100 and calibrated at 50 for the average US adult population.

### Methods

At the end of the data collection period, patients’ PHNs were used to individually match T0 and T1 survey answers. Survey answers that couldn’t be matched (missing or divergent PHN) were removed from the database. In the same way, missing data either at T0 or T1 for any given score led to the removal of those responses in the computation of mean scores and differences. Scores were computed in Microsoft Excel.

**There were 437 usable T0 surveys and 254 matched and usable T1 surveys (see**

INSERT Figure 5 in appendix). The representativeness of the survey respondents as compared to the population of the clinics’ catchment areas as well as to the clinics’ rosters was analyzed at the end of T0 data collection and results have been published elsewhere (28). Representativeness was generally good. At the end of the data collection period, we used descriptive and inferential statistics on socio-demographic information to assess the risk of attrition bias. For categorical data we used Chi square tests based on the actual T1 number of respondents per category and expected T1 numbers based on T0 proportions. Age was assessed both as a categorical variable based on 10-year age groups and by a t-test of the mean.

Primary outcomes were analyzed using paired t-tests of the mean (T1-T0) on each of the five scores related to the core dimensions of patients’ primary care experiences as well as for the mental and physical health scores. Computations were done in IBM SPSS (v26).

## RESULTS

The overall completion rate of the T1 survey from T0 respondents was 58% (individual clinics rate varied from 44% to 72%). The Chi2 tests conducted showed no significant indication of an attrition bias in the core socio-demographic variables captured by the survey (See Table 1 in appendix)

The main finding from the longitudinal analysis of patients’ experience of care is that there was a large increase in the scores for all dimensions measured (see INSERT Figure 1 below). Patient perceptions of accessibility, continuity, comprehensiveness, responsiveness and that the care received made a difference on their health were significantly higher compared to the period preceding their rostering in the clinic. Probabilities from the paired t-test of the mean are below 0.000 for all five dimensions (see Table 2 in appendix)

**Figure 1.**
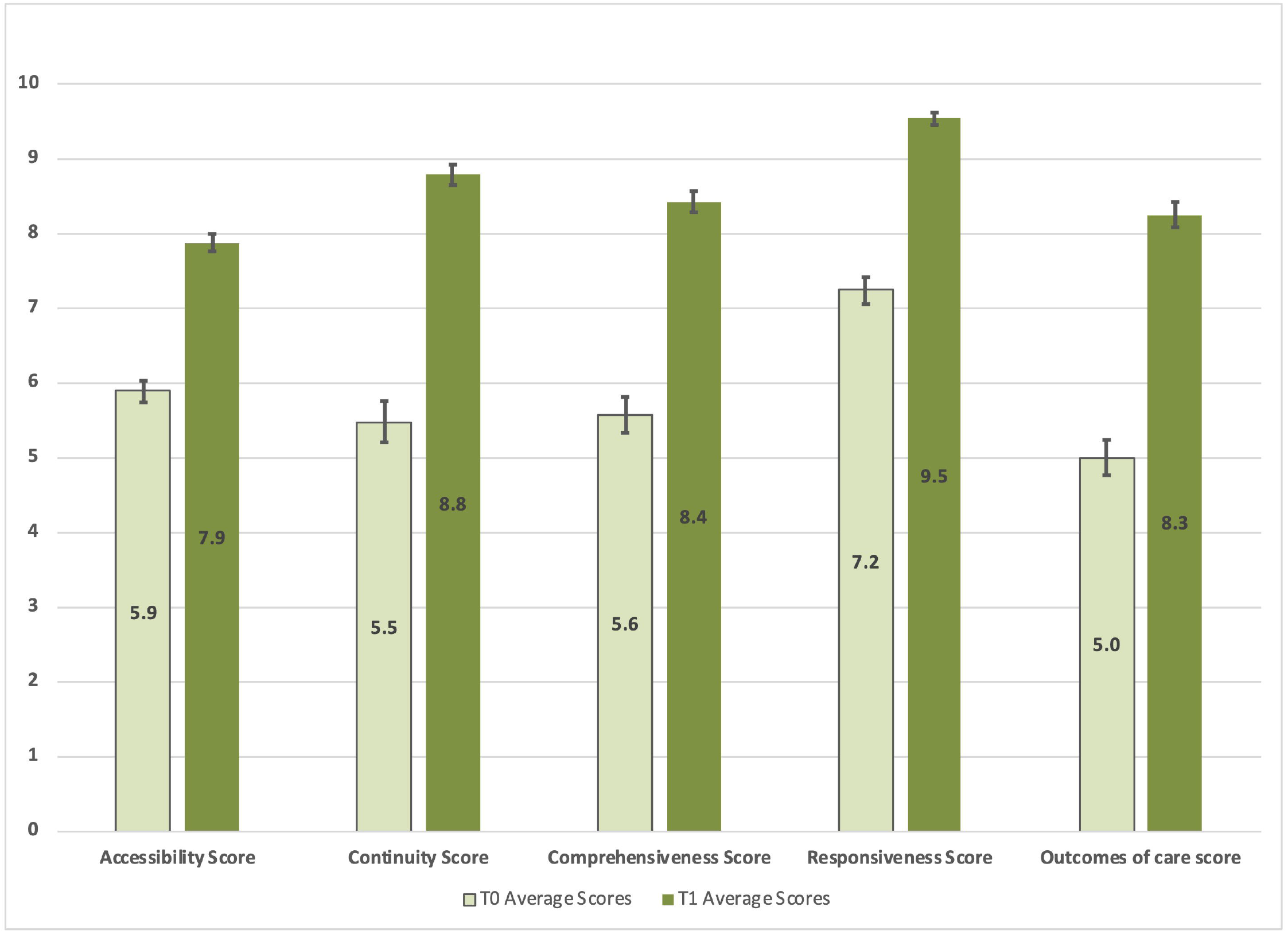
Patient Experience of Care Scores (Error bars = 95% CI)

There was some inter-site variability in pre-and post-scores but the overall trend of significant improvements remains (see INSERT Figure 2 below). All dimensions of patients’ primary care experiences vastly improved at all four sites. The pre-post differences for individual clinics are again significant (p<0.00) for all dimensions (see Table 2 in appendix).

**Figure 2.**
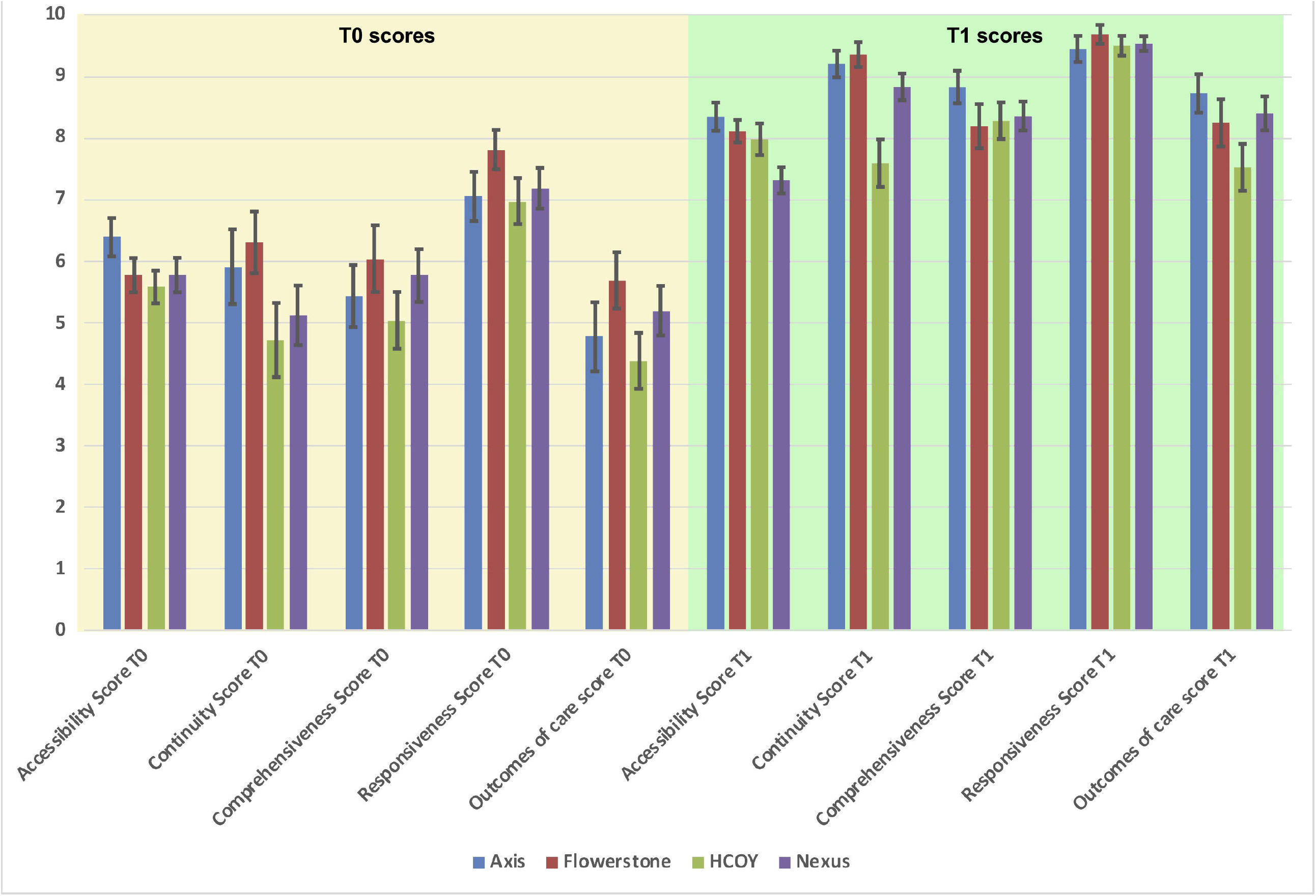
Patient Experience of Care Scores per Clinic (Error bars = 95% CI)

The composite experience of care scores also aligns with individual survey questions focused on similar constructs. For example, the proportion of respondents reporting unmet needs (based on the question, “*In the last year, did you feel the need to see a doctor or another health professional for a health problem, but didn’t see one*”) sharply decreased between T0 and T1. The overall proportion of patients reporting unmet needs at T0 was 60% (ranging from 45% at Axis to 67% at HCOY). At T1 that proportion was down to 19% (ranging from 4% at Axis to 33% at HCOY). As the question is not specific to primary care, challenges in access to specialist physicians or other services in each area should be considered in the interpretation of those scores.

Regarding respondents’ health condition as measured by the SF-12 instruments, our results show an overall improvement in the physical health scores (rising from 44.8 at T0 to 47.6 at T1 p<0.000) and no change in the mental health score (from 45.8 at T0 to 46.3 at T1 p=0.709) (see INSERT Figure 3 below as well as Table 3 in appendix).

**Figure 3.**
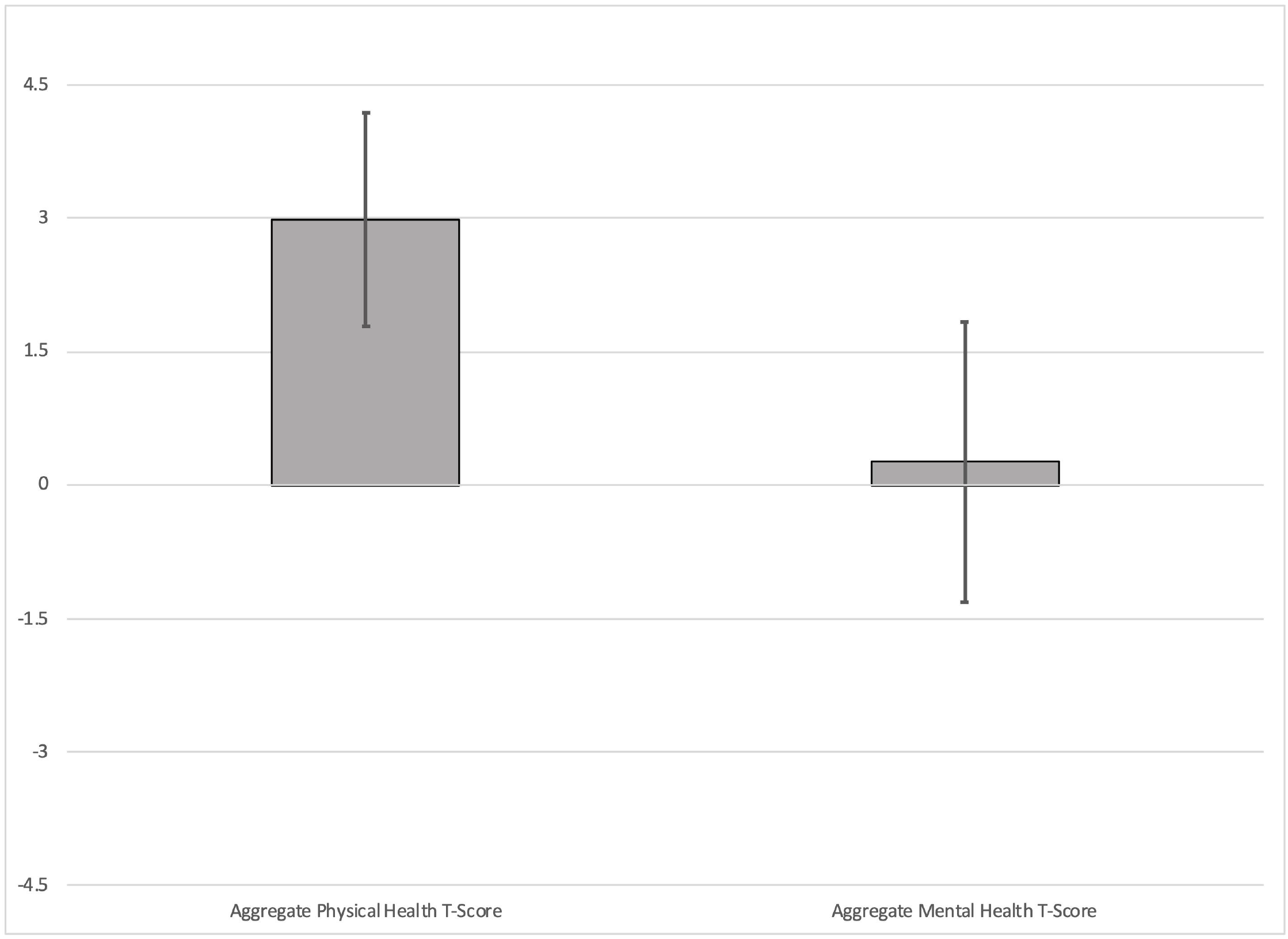
Differences in SF-12 Scores (T1-T0) Error bars = 95% CI.

**Figure 4.**
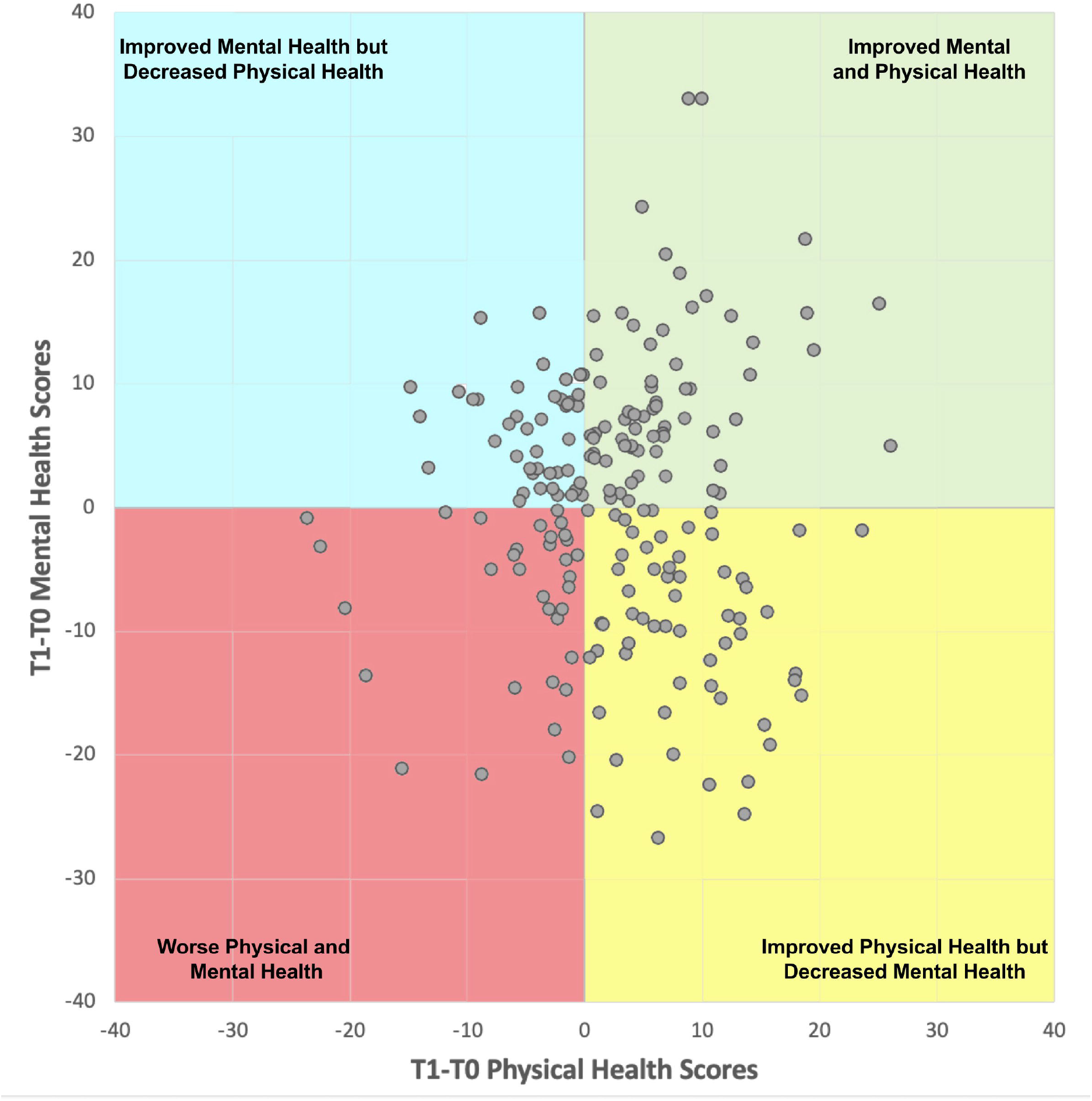
T1-T0 differences in SF-12 scores scatterplot.

When analyzed on a site-by-site basis, the trend of a significant improvement in the SF-12 physical health score persists. The data shows a significant difference (at alpha 0.05) in pre-post physical health scores at three of the four clinics. Given the low number of responses (df ranging from 32 to 69), this trend is interesting.

When the matched T1-T0 differences in physical health scores are plotted in relation to the T1-T0 differences in mental health scores, it creates a scatter plot with four quadrants. The upper right quadrant contains respondents who reported both improved physical and mental health. The lower left quadrant contains respondents who reported both decreased physical and mental health. The two other quadrants correspond to people reporting improvements in one area and a decrease in the other. Overall, the figure shows that the improvement in the average physical health score is a product of both more people on the right on the graph (62% of respondents reported an improvement in their physical health) and the fact their average score difference is higher in absolute value (+7.6) as compared to respondents with a negative physical health score difference (−5.1). No such pattern exists for the mental health score differences (top versus bottom of the figure)

## DISCUSSION

The main finding from the longitudinal analysis of patients’ experience of care is that there was a large increase in the scores for all dimensions measured after rostering in a NP-PCC. The improvements observed in patients’ experiences regarding primary care can be in part explained by the low pre-rostering (T0) baseline. Other studies that relied on the same instruments (17, 24) measured higher baseline or control-arm scores. However, the post-rostering scores (T1) measured here are similar to what was found for well-established medical clinics in those studies. Both the magnitude of the improvements measured here and the comparison of scores from the NP-PCC model as compared with the ones of large well-established interdisciplinary clinics found in other studies (17) suggest that NP-PCCs are an effective model of primary care delivery.

Given the short duration of the longitudinal follow-up conducted here, we were surprised to observe a significant improvement in the physical health as measured by the SF-12 instrument. Informal discussions with NPs working at the HCOY clinic suggest that the NP-PCCs rostered a large proportion of patients who had been left with unmet need for long periods of time. Our hypothesis is that providing accessible and comprehensive care to those patients might have been enough to cause the observed improvements in physical health. It could also be that, given recovery from mental health condition is usually longer, our study period was too short to catch improvements at that level.

More generally, it might be stressed that the NP-PCC model was, from the onset, based on the assumption those clinics would focus on rostering patients who do not have a regular provider. The survey data confirms most patients did fit that profile and that they faced low accessibility to care as well as generally low quality of care before joining the NP-PCC (28). The proportion of survey respondents reporting unmet needs at T0 (60%) is many times higher than what is observed in other populations. In 2022, 7.7 % of people in British Columbia and 7.2 Canada declared they didn’t get care when they had needs (29) while the European Union average for medical care was at 4.8% (30).

### Limitations

This study has some limitations. First, it rests on a quasi-experimental design based on a pre-post survey without a control group. As such there is a risk that external factors explain the differences observed here. The most notable external factor is that the data collection took place between 2020 and 2022, during the Covid-19 pandemic. To control for some biases, the pre-rostering survey questions were edited to ask respondents about the care they received during the “last two years” therefore including one pre-pandemic year. This phrasing is likely to have overinflated T0 scores as compared to what they would have been if those patients had to seek care outside of the NP-PCCs at that time. It might also be worth mentioning that all available indicators suggest that the primary care delivery capacity in BC sharply declined throughout the data collection period (31). Both the phrasing of the T0 survey and the context during data collection are likely to have had a negative impact on T1 to T0 score differences making the magnitude of the differences observed here even more interesting. Finally, we want to mention that the limited number of respondents is explained mostly by the fact only four NP-PCC clinics exist and that participation in the survey was voluntary and uncompensated. Nevertheless, respondents were generally representative of the total clinic roster population (28).

## CONCLUSION

Our findings show that offering people who were relying on low-continuity, low comprehensiveness services, like medical walk-in clinics, access to an NP-Led alternative dramatically improves their care experience. All dimensions of care are positively affected, and our results suggest that being followed in an NP-PCC could lead to gains in patients’ overall physical health. The patients’ primary care experience scores measured in our study favourably compare to the ones measured in well-established interdisciplinary and medical clinics.

Our findings are well aligned with the large body of evidence showing that NP-led primary care is equivalent or better than average available medical care (6, 8, 11, 32). Our study also shows that BC’s NP-PCC model does produce the results that were hoped for when the model was launched (15) and constitutes an effective approach to improve primary care accessibility and quality. As the first four NP-PCC were funded as pilots of a potential scaling up of the model, our results can play a role in future policy decisions to that effect.

**This work was supported**, in part, by the non-profit Nurses and Nurse Practitioners of British Columbia (NNPBC) association

## Data Availability

The nature of the data and conditions set by the University of Victoria Human Research Ethics Board prevent the sharing of the raw data.

## Authors contributions

DC, KB and AD designed the study. DC, KB and GR were involved in data collection. DC ran the computations and analyses and wrote the first draft. DC KB, AD and GR were involved in writing the final manuscript. The authors also want to thank Breanna Horne (Uvic), Ashleigh Swanson (Uvic), Shawna Glassel (NNPBC), Harjot Grewal (NNPBC), Lynn Guengerich (Health Care on Yates), Kari Jonker (Nexus), Lexi Grisdale (Axis) and Liz Gilmour (Flowerstone) for their support in this project.

## APPENDIX

**Figure 5.**
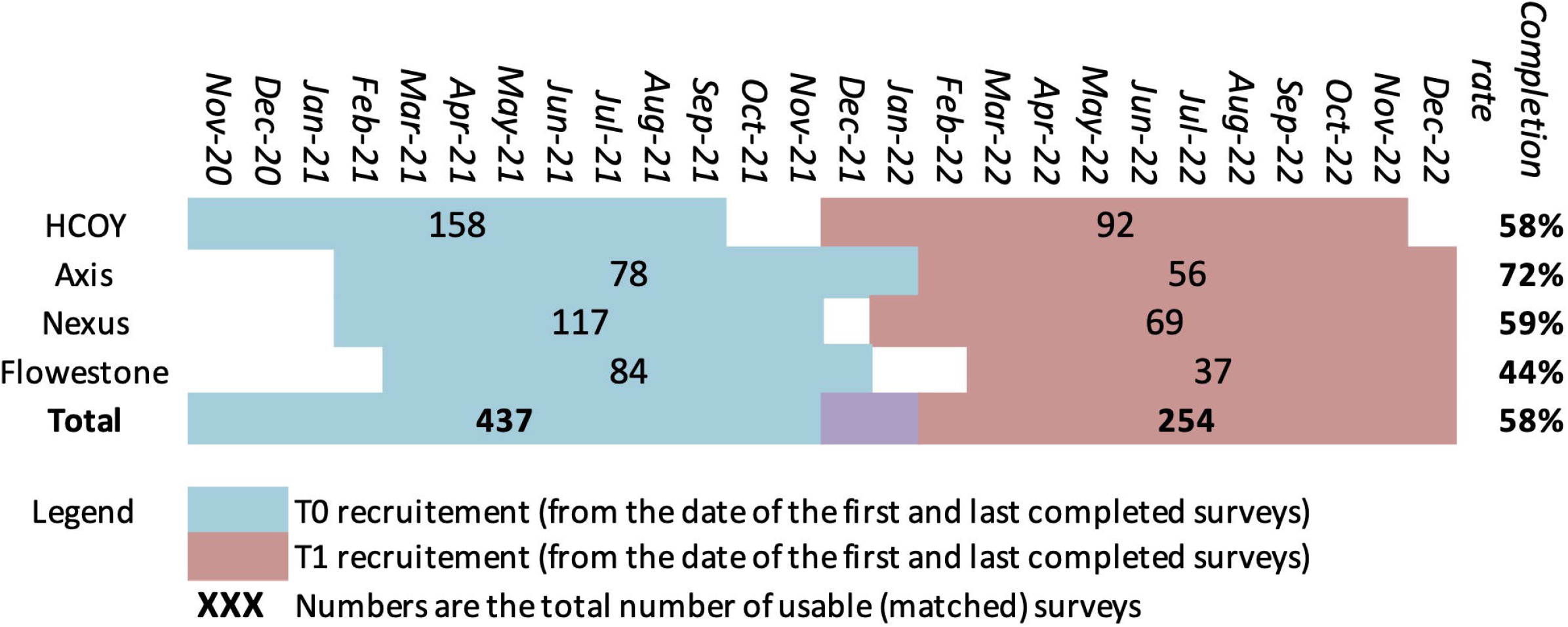
Recruitment periods, number of participants and completion rates per sites.

**Table 1.** Assessment of T0 to T1 Attrition Bias.

|  | Chi2 test probability |
| --- | --- |
| Gender (Men, Women, Other) | 0.126 |
| Age groups (10 years groups) | 0.491 |
| Born in Canada (Yes, No) | 0.086 |
| Marital status (Single, Married, Divorced or separated, Widowed) | 0.203 |
| Highest level of education (6 groups) | 0.108 |
| Occupation (8 groups) | 0.080 |
| Income (7 groups) | 0.647 |
| **t-test probability** |  |
| Mean age | 0.316 |

**Table 2.** Pre-Post Analysis of Patient Experience of Care.

| Paired Samples Test |  | Paired Differences |  |  |  |  | t | df | Sig. (2-tailed) |
| --- | --- | --- | --- | --- | --- | --- | --- | --- | --- |
|  |  | Mean | Std. Deviation | Std. Error Mean | 95% Confidence Interval of the Difference |  |  |  |  |
|  |  |  |  |  | Lower | Upper |  |  |  |
| Clinic: Axis |  |  |  |  |  |  |  |  |  |
| Pair 1 | Accessibility Score T1 - T0 | 1.959 | 2.313 | 0.353 | 1.247 | 2.671 | 5.554 | 42 | 0.000 |
| Pair 2 | Continuity Score T1 - T0 | 3.292 | 3.233 | 0.563 | 2.145 | 4.438 | 5.849 | 32 | 0.000 |
| Pair 3 | Comprehensiveness Score T1 - T0 | 3.395 | 3.200 | 0.494 | 2.398 | 4.392 | 6.875 | 41 | 0.000 |
| Pair 4 | Responsiveness Score T1 - T0 | 2.395 | 2.498 | 0.385 | 1.616 | 3.173 | 6.213 | 41 | 0.000 |
| Pair 5 | Outcomes of care score T1 - T0 | 3.953 | 3.569 | 0.551 | 2.841 | 5.065 | 7.178 | 41 | 0.000 |
| Clinic: Flowerstone |  |  |  |  |  |  |  |  |  |
| Pair 1 | Accessibility Score T1 - T0 | 2.339 | 1.950 | 0.330 | 1.669 | 3.009 | 7.096 | 34 | 0.000 |
| Pair 2 | Continuity Score T1 - T0 | 3.048 | 2.772 | 0.498 | 2.031 | 4.065 | 6.123 | 30 | 0.000 |
| Pair 3 | Comprehensiveness Score T1 - T0 | 2.153 | 3.810 | 0.663 | 0.803 | 3.504 | 3.247 | 32 | 0.003 |
| Pair 4 | Responsiveness Score T1 - T0 | 1.874 | 2.251 | 0.381 | 1.100 | 2.647 | 4.924 | 34 | 0.000 |
| Pair 5 | Outcomes of care score T1 - T0 | 2.560 | 3.551 | 0.609 | 1.320 | 3.799 | 4.203 | 33 | 0.000 |
| Clinic: HCOY |  |  |  |  |  |  |  |  |  |
| Pair 1 | Accessibility Score T1 - T0 | 2.399 | 1.959 | 0.322 | 1.746 | 3.052 | 7.450 | 36 | 0.000 |
| Pair 2 | Continuity Score T1 - T0 | 2.872 | 3.895 | 0.736 | 1.362 | 4.382 | 3.901 | 27 | 0.001 |
| Pair 3 | Comprehensiveness Score T1 - T0 | 3.244 | 2.818 | 0.463 | 2.305 | 4.184 | 7.003 | 36 | 0.000 |
| Pair 4 | Responsiveness Score T1 - T0 | 2.527 | 2.199 | 0.361 | 1.794 | 3.260 | 6.990 | 36 | 0.000 |
| Pair 5 | Outcomes of care score T1 - T0 | 3.145 | 3.312 | 0.545 | 2.040 | 4.249 | 5.775 | 36 | 0.000 |
| Clinic: Nexus |  |  |  |  |  |  |  |  |  |
| Pair 1 | Accessibility Score T1 - T0 | 1.539 | 2.181 | 0.294 | 0.949 | 2.128 | 5.233 | 54 | 0.000 |
| Pair 2 | Continuity Score T1 - T0 | 3.714 | 3.355 | 0.479 | 2.750 | 4.677 | 7.748 | 48 | 0.000 |
| Pair 3 | Comprehensiveness Score T1 - T0 | 2.589 | 3.512 | 0.478 | 1.631 | 3.548 | 5.417 | 53 | 0.000 |
| Pair 4 | Responsiveness Score T1 - T0 | 2.352 | 2.424 | 0.330 | 1.690 | 3.014 | 7.129 | 53 | 0.000 |
| Pair 5 | Outcomes of care score T1 - T0 | 3.206 | 3.486 | 0.483 | 2.236 | 4.177 | 6.632 | 51 | 0.000 |
| Total (All Clinics) |  |  |  |  |  |  |  |  |  |
| Pair 1 | Accessibility Score T1 - T0 | 1.997 | 2.135 | 0.164 | 1.674 | 2.320 | 12.194 | 169 | 0.000 |
| Pair 2 | Continuity Score T1 - T0 | 3.301 | 3.306 | 0.278 | 2.751 | 3.852 | 11.857 | 140 | 0.000 |
| Pair 3 | Comprehensiveness Score T1 - T0 | 2.852 | 3.359 | 0.261 | 2.338 | 3.367 | 10.940 | 165 | 0.000 |
| Pair 4 | Responsiveness Score T1 - T0 | 2.301 | 2.350 | 0.181 | 1.943 | 2.659 | 12.692 | 167 | 0.000 |
| Pair 5 | Outcomes of care | 3.249 | 3.484 | 0.271 | 2.714 | 3.785 | 11.980 | 164 | 0.000 |
| 5 | score T1 - T0 |  |  |  |  |  |  |  |  |

**Table 3.** Pre-Post Analysis of Health (SF-12) Improvements.

| Paired Samples Test |  | Paired Differences |  |  |  |  | t | df | Sig. (2-tailed) |
| --- | --- | --- | --- | --- | --- | --- | --- | --- | --- |
|  |  | Mean | Std. Deviation | Std. Error Mean | 95% Confidence Interval of the Difference |  |  |  |  |
|  |  |  |  |  | Lower | Upper |  |  |  |
| Clinic: Axis |  |  |  |  |  |  |  |  |  |
| Pair 1 | Aggregate Physical Health T-Score (T1-T0) | 2.721 | 8.054 | 1.243 | 0.211 | 5.230 | 2.189 | 41 | 0.034 |
| Pair 2 | Aggregate Mental Health T-Score (T1-T0) | -0.752 | 9.893 | 1.527 | -3.835 | 2.331 | -0.493 | 41 | 0.625 |
| Clinic: Flowerstone |  |  |  |  |  |  |  |  |  |
| Pair 1 | Aggregate Physical Health T-Score (T1-T0) | 4.060 | 7.917 | 1.378 | 1.253 | 6.867 | 2.946 | 32 | 0.006 |
| Pair 2 | Aggregate Mental Health T-Score (T1-T0) | 3.806 | 8.750 | 1.523 | 0.703 | 6.908 | 2.498 | 32 | 0.018 |
| Clinic: HCOY |  |  |  |  |  |  |  |  |  |
| Pair 1 | Aggregate Physical Health T-Score (T1-T0) | 1.770 | 7.696 | 0.920 | -0.065 | 3.605 | 1.924 | 69 | 0.058 |
| Pair 2 | Aggregate Mental Health T-Score (T1-T0) | -1.249 | 11.581 | 1.384 | -4.010 | 1.512 | -0.902 | 69 | 0.370 |
| Clinic: Nexus |  |  |  |  |  |  |  |  |  |
| Pair 1 | Aggregate Physical Health T-Score (T1-T0) | 3.446 | 9.026 | 1.175 | 1.094 | 5.799 | 2.933 | 58 | 0.005 |
| Pair 2 | Aggregate Mental Health T-Score (T1-T0) | 0.849 | 10.595 | 1.379 | -1.912 | 3.610 | 0.616 | 58 | 0.541 |
| Total (All Clinics) |  |  |  |  |  |  |  |  |  |
| Pair 1 | Aggregate Physical Health T-Score (T1) - (T0) | 2.822 | 8.193 | 0.574 | 1.691 | 3.953 | 4.919 | 203 | 0.000 |
| Pair 2 | Aggregate Mental Health T-Score (T1) - (T0) | 0.277 | 10.613 | 0.743 | -1.188 | 1.742 | 0.373 | 203 | 0.709 |

## REFERENCES

1. Statistics Canada. Primary health care providers, 2019. Ottawa: Minister of Industry; 2020.

2. Commonwealth Fund survey, 2020 [Internet]. CIHI. 2020. Available from: https://www.cihi.ca/en/commonwealth-fund-survey-2020.

3. Contandriopoulos D, Brousselle A, Breton M, Sangster-Gormley E, Kilpatrick K, Dubois C-A, et al. Nurse practitioners, canaries in the mine of primary care reform. Health Policy. 2016;120(6):682–9.

4. Heale R, Butcher M. Canada’s first nurse practitioner-led clinic: a case study in healthcare innovation. Nurs Leadersh. 2010;23(3):21–9.

5. Lovink MH, Persoon A, Koopmans RT, Van Vught AJ, Schoonhoven L, Laurant MG. Effects of substituting nurse practitioners, physician assistants or nurses for physicians concerning healthcare for the aging population: A systematic literature review. Journal of advanced nursing. 2017;73(9):2084–102.

6. Martin-Misener R, Harbman P, Donald F, Reid K, Kilpatrick K, Carter N, et al. Cost-effectiveness of nurse practitioners in primary and specialised ambulatory care: systematic review. BMJ Open. 2015;5(6):e007167.

7. Poghosyan L, Liu J, Norful AA. Nurse practitioners as primary care providers with their own patient panels and organizational structures: A cross-sectional study. International Journal of Nursing Studies. 2017;74:1–7.

8. Laurant M, van der Biezen M, Wijers N, Watananirun K, Kontopantelis E, van Vught Ajah. Nurses as substitutes for doctors in primary care. Cochrane Database of Systematic Reviews. 2018(7):Art. No.: CD001271.

9. Christensen C, Grossman J, Hwang J. The Innovator’s Prescription: A Disruptive Solution for Health Care. New-York: McGraw Hill; 2016.

10. Contandriopoulos D, Perroux M, Cockenpot A, Duhoux A, Jean E. Analytical typology of multiprofessionnal primary care models. BMC Family Practice. 2018;19(44):1–11.

11. Greene J. Nurse practitioners provide quality primary care at a lower cost than physicians. Manag Care. 2018;27(11):34.

12. Liu CF, Hebert PL, Douglas JH, Neely EL, Sulc CA, Reddy A, et al. Outcomes of primary care delivery by nurse practitioners: Utilization, cost, and quality of care. Health Serv Res. 2020;55(2):178–89.

13. Randall S, Crawford T, Currie J, River J, Betihavas V. Impact of community based nurse-led clinics on patient outcomes, patient satisfaction, patient access and cost effectiveness: A systematic review. International Journal of Nursing Studies. 2017;73:24–33.

14. Prodan-Bhalla N, Scott L. Primary Care Transformation in British Columbia: A New Model to Integrate Nurse Practitioners. Vancouver: British Columbia Nurse Practitionner Association; 2016.

15. British Columbia Ministry of Health. New nurse practitioner primary care clinic opening soon in Victoria 2020 [Available from: https://news.gov.bc.ca/releases/2020HLTH0296-001789.

16. Contandriopoulos D, Duhoux A, Roy B, Amar M, Bonin J-P, Borges Da Silva R, et al. Integrated Primary Care Teams (IPCT) pilot project in Quebec: a protocol paper. BMJ Open. 2015;5(e010559).

17. Duhoux A, Dufour É, Sasseville M, Laroche D, Contandriopoulos D. Rethinking Primary Care Delivery Models: Can Integrated Primary Care Teams Improve Care Experience? International Journal of Integrated Care. 2022;22(8):1–12.

18. Haggerty JL, Burge F, Beaulieu M-D, Pineault R, Beaulieu C, Lévesque J-F, et al. Validation of Instruments to Evaluate Primary Healthcare from the Patient Perspective: Overview of the Method. Healthcare Policy. 2011;7(Special Issue):31–46.

19. Levesque J-F, Descôteaux S, Demers N, Benigeri M. Measuring Organizational Attributes of Primary Healthcare: A Scanning Study of Measurement Items Used in International Questionnaires Montréal: Unité Évaluation de l’organisation des soins et services, Direction de l’analyse et de l’évaluation des systèmes de soins et services, INSPQ; 2014.

20. Pineault R, Levesque J-F, Roberge D, Hamel M, Lamarche P, Haggerty J. L’accessibilité et la continuité des services de santé : une étude sur la première ligne au Québec : rapport de recherche. Longueuil: Centre de Recherche de l’Hôpital Charles LeMoyne; 2008.

21. Donabedian A. The Quality of Care. How can it be assessed? Journal of the American Medical Association. 1988;206(12).

22. Haggerty JL, Burge F, Pineault R, Beaulieu M-D, Bouharaoui F, Beaulieu C, et al. Management Continuity from the Patient Perspective: Comparison of Primary Healthcare Evaluation Instruments. Healthcare Policy. 2011;7(Special Issue):139–53.

23. Haggerty JL, Beaulieu M-D, Pineault R, Burge F, Lévesque J-F, Santor DA, et al. Comprehensiveness of Care from the Patient Perspective: Comparison of Primary Healthcare Evaluation Instruments. Healthcare Policy. 2011;7(Special Issue):154–66.

24. Pineault R, Silva RBD, Provost S, Breton M, Tousignant P, Fournier M, et al. Impacts of Québec primary healthcare reforms on patients’ experience of care, unmet needs, and use of services. International Journal of Family Medicine. 2016:Epub.

25. Haidar OM, Lamarche PA, Levesque J-F, Pampalon R. The Influence of Individuals’ Vulnerabilities and Their Interactions on the Assessment of a Primary Care Experience. International Journal of Health Services. 2018;48(4):798–819.

26. Safran DG, Kosinski M, Tarlov AR, Rogers WH, Taira DH, Lieberman N, et al. The Primary Care Assessment survey: Tests of data quality and measurement Performance. Medical Care. 1998;36(5):728–39.

27. Shi L, Starfield B, Xu J. Validating the Adult Primary Care Assessment Tool. Journal of Family Practice. 2001;50(2):161–71.

28. Contandriopoulos D, Bertoni K, Horne B, Swanson A, Glassel S, Guengerich L, et al. A descriptive analysis of the previous care experiences of patients being rostered in BC’s new Nurse-Practitioner Primary Care Clinics. Nurse Practitioner Open Journal. 2023;In press.

29. Statistics Canada. Table 13-10-0836-01 Unmet health care needs by sex and age group: Statistics Canada; 2022 [Available from: https://www150-statcan-gc-ca.ezproxy.library.uvic.ca/t1/tbl1/en/tv.action?pid=1310083601.

30. Eurostat. Unmet health care needs statistics (2022): Eurostat.; 2022 [Available from: https://ec.europa.eu/eurostat/statistics-explained/index.php?title=Unmet_health_care_needs_statistics#Unmet_needs_for_health_care.

31. Xu X. Nearly 900,000 British Columbians don’t have a family doctor, leaving walk-in clinics and ERs swamped. The Globe and Mail. 2022 April 29, 2022.

32. Mundinger MO, Kane RL, Lenz ER, Totten AM, Tsai W-Y, Cleary PD, et al. Primary Care Outcomes in Patients Treated by Nurse Practitioners or Physicians: A Randomized Trial. JAMA. 2000;283(1):59–68.

33. Wilson, E. C., Pammett, R., McKenzie, F., & Bourque, H. (2021, Jan-Mar). Engagement of nurse practitioners in primary health care in northern British Columbia: a mixed-methods study. CMAJ Open, 9(1), E288–e294. https://doi.org/10.9778/cmajo.20200075

